# Automated Bias Reduction in Deep Learning Based Melanoma Diagnosis using a Semi-Supervised Algorithm

**DOI:** 10.1101/2021.01.13.21249774

**Authors:** Sauman Das

## Abstract

Melanoma is one of the most fatal forms of skin cancer and is often very difficult to differentiate from other benign skin lesions. However, if detected at its early stages, it can almost always be cured. Researchers and data scientists have studied this disease in-depth with the help of large datasets containing high-quality dermascopic images, such as those assembled by the International Skin Imaging Collaboration (ISIC). However, these images often lack diversity and over-represent patients with very common skin features such as light skin and having no visible body hair. In this study, we introduce a novel architecture called *LatentNet* which automatically detects over-represented features and reduces their weights during training. We tested our model on four distinct categories - three skin color levels corresponding to Type I, II, and III on the Fitzpatrick Scale, and images containing visible hair. We then compared the accuracy against the conventional Deep Convolutional Neural Network (DCNN) model trained using the standard approach (i.e. without detecting over-represented features) and containing the same hyper-parameters as the LatentNet. LatentNet showed significant performance improvement over the standard DCNN model with accuracy of 89.52%, 79.05%, 64.31%, and 64.35% compared to the DCNN accuracy of 90.41%, 70.82%, 45.28%, 56.52% in the corresponding categories, respectively. Differences in the average performance between the models were statistically significant (*p* < 0.05), suggesting that the proposed model successfully reduced bias amongst the tested categories. LatentNet is the first architecture that addresses racial bias (and other sources of bias) in deep-learning based Melanoma diagnosis.

## 1. Introduction

Melanoma is one of the deadliest forms of skin cancer and makes up 75% of deaths related to skin cancer (Li and Shen, 2018). According to American Cancer Society, each year, roughly 100,350 people are diagnosed with Melanoma resulting in 6,850 deaths in the USA. Although Melanoma is a malignant form of skin cancer, it can be treated with almost 100% survival rate if diagnosed at its early stages (Friedman et al., 1985).

Melanoma occurs when *melanocytes*, pigment-producing cells become cancerous. The thickness of the lesion grows as the disease progresses. Friedman et al. (1985) showed that the disease yields a 10-year survival rate of 99.5% if diagnosed when the thickness of the lesion is under 0.75mm. However, once the thickness grows over 3.00mm, the 10-year survival rate plummets to 48%. Therefore, diagnosing Melanoma in early stages is essential in maximizing the chances of patient’s survival.

Typically medical professionals diagnose Melanoma by examining a tissue sample from a patient’s skin lesion. However, studies have shown that the diagnosis through analysis of histologic features often varies based on the experience and skill level of the medical observer (Scolyer et al., 2011). To overcome this issue, recent research has been focused on developing more objective approaches of diagnosing Melanoma.

*Dermoscopy* was developed to provide more accurate results for diagnosing skin lesions. It is a noninvasive imaging technique that magnifies a small skin lesion as shown in Figure 1 (Binder et al., 1995). Although the dermoscopic imaging technique was initially created to help dermatologists diagnose skin lesions, these images are commonly used to train deep learning models to exploit the detailed features in the images. Due to the availability of large dermoscopic images over the past decade, deep learning approaches have yielded promising results. These machine learning based automatic systems are trained on thousands of these images.

**Figure 1:**
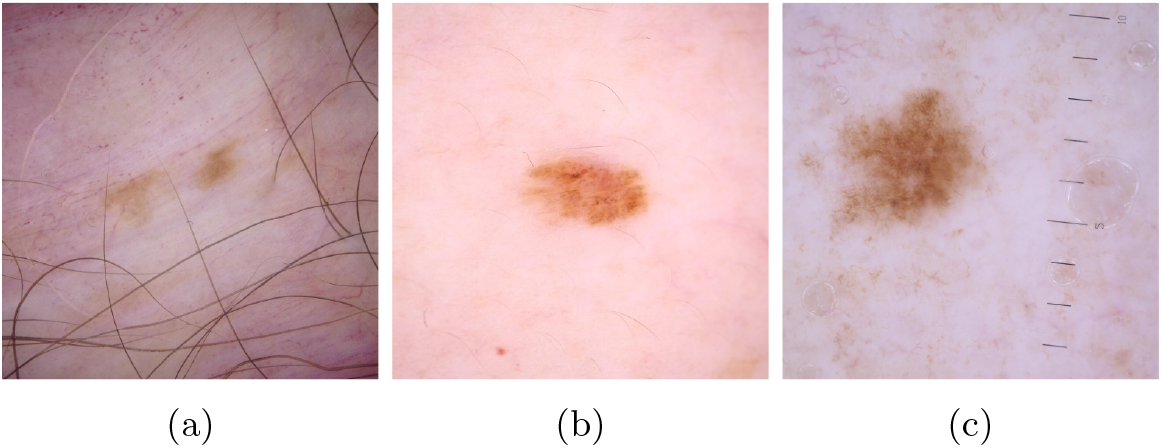
Sample dermascopic images shown.

While current state-of-the-art models can achieve high diagnosis accuracy, they are mostly trained and evaluated on images from the same source. However, every dataset contains some inherent bias, typically over representing certain features more than others. For example, a dataset may contain more images without hair than with hair. The model trained on this dataset will have a bias in the sense that the diagnosis accuracy will be higher for images without hair compared to the the images without hair.

In this paper, given the a dataset, we attempt to create a model that will improve diagnosis accuracy across several variations of physical characteristics including skin color levels and images with the presence of hair. We will first investigate previous research about how Melanoma has been previously solved using deep learning along with other articles that discuss varying approaches to countering bias in Section 2. Then, in Section 3, we build two models, one trained using a general approach from previous papers, and the other trained intelligently to reduce bias. In Section 4, we discuss our results along with details about how we went about evaluating bias. Finally, Section 5 outlines our conclusions and ideas to build upon this research in the future.

## 2. Background

Several papers have been published on providing an objective diagnosis for Melanoma using deep learning. Each project has taken a different approach to achieving this goal. In Li and Shen (2018) the authors utilized a Deep Convolutional Neural Network (DCNN) model to achieve an overall 91.2% accuracy when classifying dermascopic images. Their custom DCNN model was an important part of the process to extract relevant features from the images before producing an output/diagnosis. In Codella et al. (2015) the authors followed a similar DCNN approach to perform feature extraction. However, they used a Support Vector Machine (SVM) model after feature extraction to achieve a 93.1% overall accuracy.

For performance evaluation, both of the above papers utilized dermascopic images from the International Skin Imaging Collaboration (ISIC). ISIC is known to be the largest open-source database for labelled der-mascopic images and has been adding to the archive of images over the years and recently partnered with the Society for Imaging Informatics in Medicine (SIIM) to update the dataset (Tschandl et al., 2018). It is stated that the images for ISIC’s HAM10000 dataset are collected from Barcelona (Spain), New York City (United States), Wollstonecraft (Australia), Brisbane (Australia), and Athens (Greece) (International Skin Imaging Collaboration, 2020).

One can imagine capturing new images to enhance the existing image dataset to remove the imbalance towards under-represented features. However, this could be a very challenging task as it took more than twenty years to collect and label the images in the HAM10000 dataset published on ISIC (Tschandl et al., 2018). Fortunately, one can learn from research in other fields to address similar dataset bias. One approach is to generate completely new images from the existing training data to fill the gap of under-represented categories. In Salehinejad et al. (2018), the authors implemented a Generative Adversarial Network (GAN) to address the scarcity of X-ray images in the rare disease categories. Using a GAN, they augmented their dataset to include artificial images which improved the DCNN model performance. GANs are a good solution to the data bias problem if the biased category can be *explicitly* defined. For example, Salehinejad et al. (2018) found that their dataset was biased towards healthy X-ray scans. Therefore, they produced more images of X-rays where the patient showed unhealthy symptoms.

Amini et al. (2019) had developed a method to detect bias for an image recognition program which differentiated faces from other arbitrary objects. They implemented a Variational Autoencoder (VAE) to automatically detect over-represented features and reduce their weight during training. In contrast to GANs, they did not define a specific category to reduce bias in. Instead, common image features were automatically detected and their weights were adjusted downwards during training to reduce bias.

Our proposed approach adapts the VAE method for melanoma diagnosis. This is a logical choice since specific skin features which are under- or over-represented are not known *a priori*. In other words, the model should automatically determine these features to reduce bias. Next we discuss two separate models. First, we build and train a DCNN model as a baseline classification system. This model is trained using the conventional approach. However, our second model named *LatentNet*, is built with the same classification structure as the DCNN, but trained using a method targeted towards reducing bias. Although we adapt the VAE method proposed by Amini et al., certain modification have been made to specifically detect features in dermascopic images rather than facial images.

## 3. Proposed Approach

### 3.1 Baseline DCNN

We first describe the fundamentals of standard DCNN model. For a good overview of convolutional neural network models, the reader can refer O’Shea and Nash (2015). A basic DCNN model as in Fig 2 extracts features from the images and produces a binary output of either 0 (benign lesion) or 1 (malignant melanoma lesion). DCNNs have proved to be a powerful tool when extracting important features from images. These types of models apply two dimensional convolutions across images to highlight important features from the input image before providing an output.

**Figure 2:**
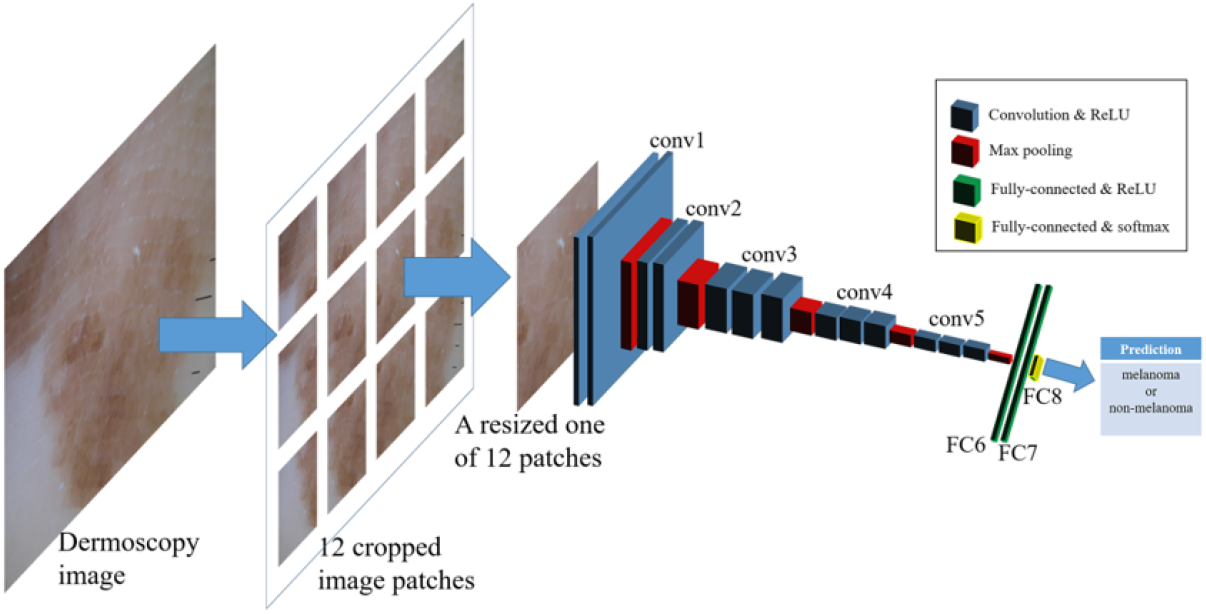
Example DCNN model. Model performs feature extraction on input image and outputs binary diagnosis (Yu et al., 2018)

For baseline comparison we used a simple sequential DCNN model as shown in Fig 3. The model consists of five convolutional layers each containing a 5×5 kernel to detect features. Each convolutional layer is followed by a batch normalization step to help train the model faster and more accurately (Ioffe and Szegedy, 2015). After the last convolutional layer, we added one more fully connected layer with 512 neurons. All layers used Rectified Linear Unit (ReLU) activation function to enable the model to learn non-linearity pattern in the data. In the final step, there is one sigmoid output neuron (*z*_0_ ∈ (0, 1)) for the binary prediction.

**Figure 3:**
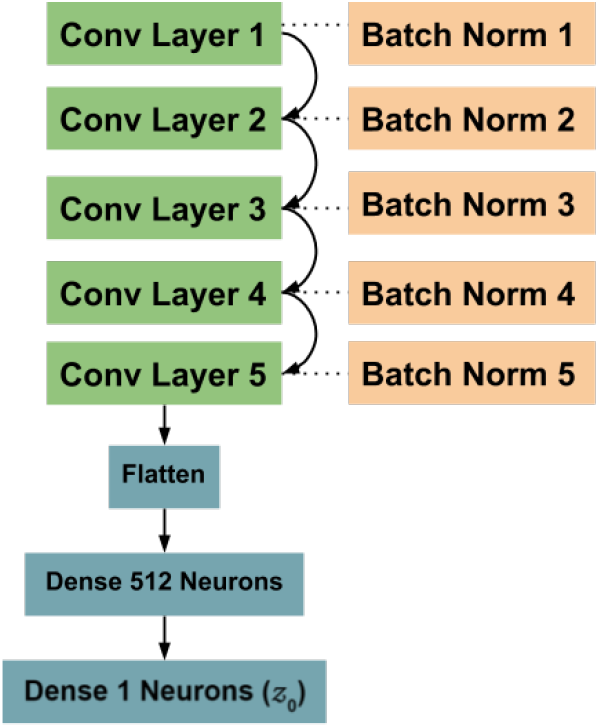
DCNN model architecture overview.

The standard model was trained with the binary cross-entropy loss function as in equation 1. Furthermore, the Adam optimizer (Kingma and Ba, 2014) was used with a learning rate of 5e-4 for 25 epochs of training.

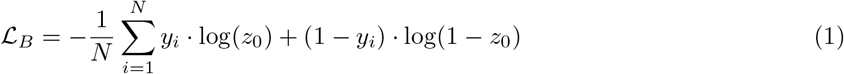

*y*_*i*_ denotes the ground truth (0 or 1) for the *i*th image and *N* specifies the number of images. The standard DCNN model was trained on 256×256×3 dermascopic images from the 2020 ISIC-SIIM dataset. The number of images was 1,168 after randomly under-sampling from the original dataset. The condensed data consisted of 584 melanoma and 584 benign images to train on.

Under-sampling is a technique we used to introduce balance into our dataset. The original 2020 ISIC-SIIM dataset contained over 20,000 dermascopic images. However, only 584 of these were Melanoma while the rest were benign lesions. If we trained our model with all of these images, then our model would always predict “No Melanoma” since it would still result in a accuracy greater than 98%. To prevent this from happening, we randomly selected 584 benign lesions to completely remove the data imbalance.

In DCNN framework, the model is trained on small batches of images selected repeatedly every training iteration. Algorithm 1 shows a simplified overview of this training process for one epoch. After initializing number of total images *n* and the batch size *b*, we repeatedly sample batches of images randomly and pass them through the DCNN producing output *z*_*batch*_ with 32 binary predictions corresponding to each image in a batch. Based on the resulting value of loss function 𝓛_*B*_, the model weights are updated through back propagation. This process is repeated every epoch.

#### Algorithm 1

DCNN Training Process for 1 Epoch

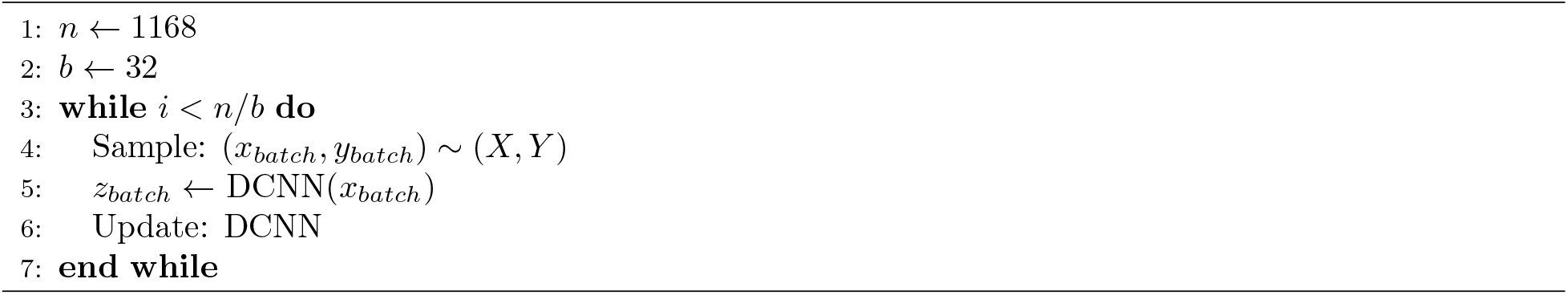

Please note that in line 4 of above batch selection algorithm, 32 labelled images are selected ***randomly*** from the full dataset of 1,168 images. Due to this purely random selection, the batch of images selected will inherit the data bias inherent in the full dataset. For example, if the full dataset has a bias towards light-skinned images, the batch will also tend to have the same bias. As a direct result of the bias in the batch selection, the DCNN based trained model will have better prediction accuracy for patients with over-represented features. In Table 1, we provide formal definitions for three levels of bias in the training process.

**Table 1:** Defining bias at three levels of the machine learning process. Data bias leads to batch bias which eventually leads to a biased model.

| Level of Bias | Definition |
| --- | --- |
| Data Bias | Training dataset does not uniformly represent all variations of skin features. |
| Batch Bias | Sample of images for each training iteration does not uniformly represent all variations of skin features. |
| Model Bias | Final model does not output accurate diagnoses for patients containing variations of skin features uncommon in the dataset. |

Although the root cause of a biased model is the bias inherent in the original dataset (Data Bias), one can fix the bias in the selected batches (Batch Bias) in order to reduce the bias in the model (Model Bias). With this concept in mind, in the following section, we will explore how to intelligently sample the batch to reduce the model bias.

### 3.2 Understanding Latent Structure

Variational Autoencoder (VAE) was introduced in the seminal paper by Kingma and Welling (2013). A machine learning model based on VAE is trained through a completely unsupervised approach. Broadly speaking, the input of the VAE is an image (*x*), and the output is a reconstruction of that same image 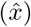 as depicted in Figure 4.

**Figure 4:**
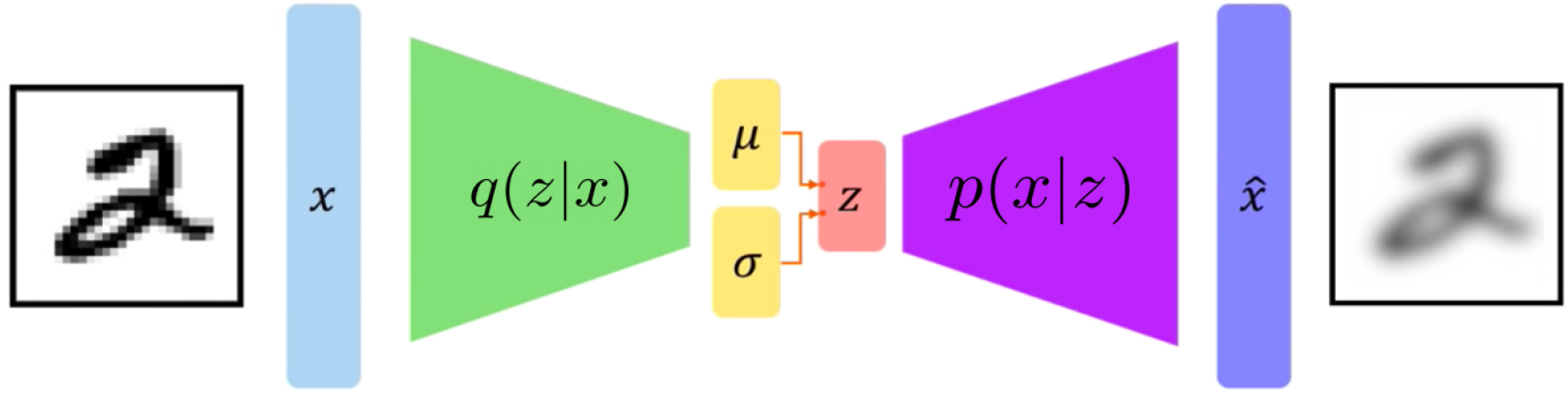
Structure of the Variational Autoencoder. Image adapted from Paul (2020).

The goal of the VAE is to create the most accurate reconstruction of the original image. Its structure contains 3 main parts: encoder, latent vector, and decoder. The encoder, *q*(*z*|*x*), is a neural network that extracts features from the input image and stores the significant features in terms of a mean vector *µ* and standard deviation vector *σ*. From the given *µ* and *σ* vectors, the final latent vector *z* is formed by sampling from a Gaussian distribution, i.e. *z* ∼ 𝒩 (*μ, σ*^2^). Given the latent vector z, the decoder, *p*(*x*|*z*), reverses the encoder to try and reconstruct the original image. The overall goal of the VAE is to store only the significant features in the compressed latent vector. As a result, the decoder will be able to interpret these features and eventually recreate the input image. For an intuitive example, assume that input *x* is a three dimensional object. The latent vector *z* can be interpreted as the shadow of the input image. A shadow approximates the original image by storing only its important features. The decoder would then look at only the shadow and try to recreate the original object. The latent structure is a valuable tool since it automatically learns only relevant features for accurate image reconstruction.

### 3.3 LatentNet Architecture

In case of a dermascopic image, the latent structure can use features such as the skin color, size of the lesion, amount of hair, etc. As discussed earlier, the benefit of the VAE lies in its ability to automatically determine these features unlike the GAN framework where classes are explicitly defined. However, a VAE alone only produces an image reconstruction without determining diagnosis based on the features. This shortcoming can be overcome easily by combining DCNN with VAE in the new proposed LatentNet architecture as depicted in Figure 5. Note that the component to the left of the Latent Space in this figure is identical to the DCNN of Figure 3.

**Figure 5:**
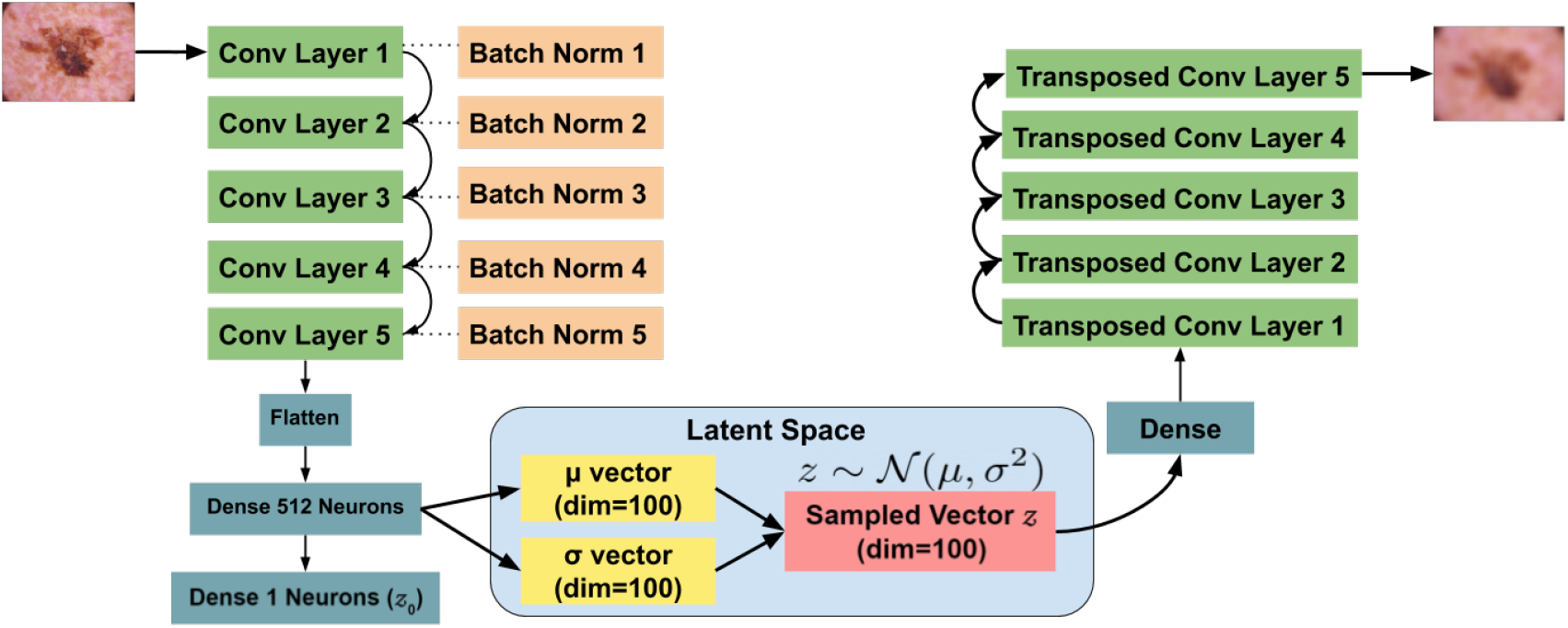
LatentNet Architecture.

The LatentNet has a encoder and a decoder just like the VAE. Looking carefully, we notice that the encoder has the exact same architecture as the DCNN except the last dense layer with 512 neurons connects to three separate layers instead of just one. It remains connected to the sigmoid output layer. However, it also connects to a *µ* and *σ* vector which lead to the sampled latent vector *z*. Note that we have set the dimension of all the latent space vectors to 100. This means that the model will compress the original 256×256×3 image pixels to just 100 numbers. The decoder “decodes” these 100 values and generates a reconstruction. The decoder is the exact opposite structure of the encoder. Each convolutional layer from the encoder is simply transposed.

The size of latent space dimensions was carefully chosen to be 100. With a very high value the model will not understand the important features. It may simply store the specific pixel values from the original image which defeats the purpose of the latent space. On the contrary, too few dimensions of the latent space means limiting to only a few features. In this scenario the reconstruction would be very poor.

### 3.4 Intelligent Batch Sampling

In LatentNet architecture, we can use the latent space vectors to intelligently sample batches of images avoiding pure random sampling. In this section, we describe how to analyze the *µ* vector from the latent space to generate probabilities of selecting a specific image for a batch.

Let *N* be the total number of images available. While training the standard DCNN model, the probability of selecting each image for a batch is 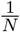. However, we want to assign images with rare variations of features to have a higher probability of selection. Consequently, the model will be able to improve the accuracy of predictions on images with these rare variations. Rare variations of skin features may include dark skin, freckles, high hair density, unusual anatomical site, etc. We now proceed to formally distinguish *rare* variations from common variations of a given feature.

Amini et al. (2019) proposed a method to assign probabilities of selecting an image for a batch based on its latent *µ* vector. The *µ* vector for each image can be obtained by passing it through the encoder. Thus, there will be *N* such *µ* vectors of length 100 corresponding to the *N* images in the dataset.

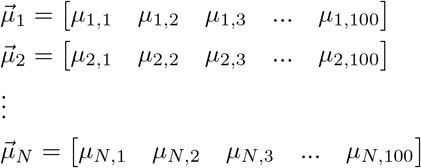

*µ*_*i,j*_ denotes the mean value for a feature *j* in the *i*th image where *i* ∈ [1, *N*] and *j* ∈ [1, 100]. We start by plotting the histogram of the *µ* values of the first latent feature corresponding to *j* = 1. All *µ* values in a specific bin of the histogram exhibit a similar variation of the corresponding feature *j*.

Figure 6 depicts an example histogram with *N* = 1, 168 images for *j* = 1. Notice that the histogram bars have a significant variation in their heights. The tallest bar represents the most common variation of feature 1. On the contrary, the shortest bar is the rarest variation of the feature. For the *i*th image, *µ*_*i*,1_ falling in the 8th bin indicates that the image exhibits the most common variation of the 1st feature. In our sampling algorithm, we give the images that lie in the 8th bin the lowest sampling probability for a batch. In contrast, we assign the few images with *µ* values falling in the shortest bin 1, the highest sampling probability since they show a rare variation of the feature. We outline the sampling process in Algorithm 2.

**Figure 6:**
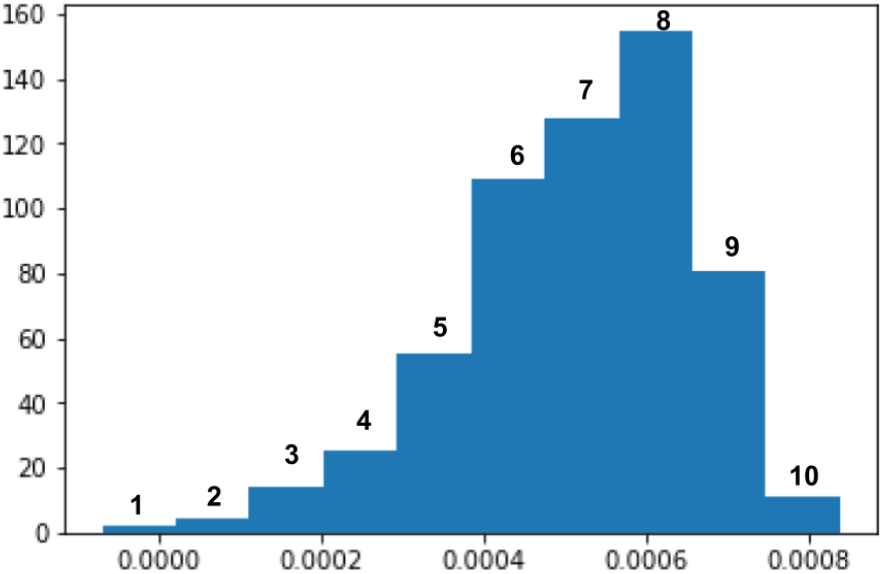
Sample histogram of 1,168 *µ* values with 10 bins. Numbers above bars represent *bin index, not frequency*.

#### Algorithm 2

Generating Sampling Probabilities

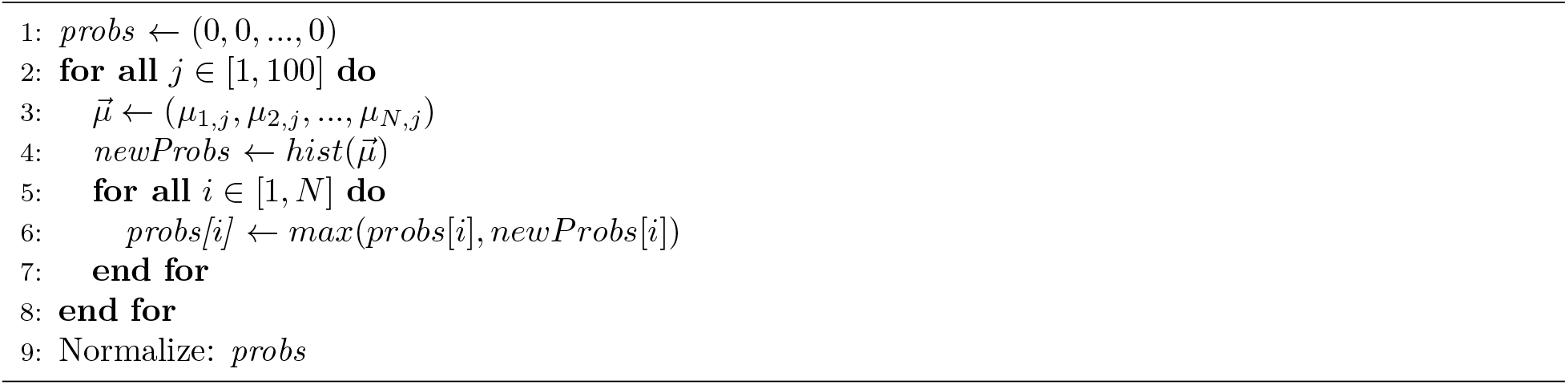

We start by initializing the probability of selecting each image as 0. The dimension of *probs* is *N*, the number of images. We iterate through all 100 latent *µ* values. At each iteration, we first create the 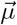 which stores the *j*th *µ* value for each image. As elaborated in the previous paragraph, we plot 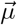 in a histogram and compute the probabilities which are inversely proportional to the height of the corresponding bin. In lines 5 through 7, probability for each image is updated by replacing the old probability, *probs*[*i*], with the new probability, *newProbs*[*i*], if and only if *newProbs*[*i*] > *probs*[*i*]. Finally, all the probability values are normalized by dividing each value by the sum of all probability values in the *probs* array. After normalization, the sum of all the probabilities in *probs* add to 1. We use this algorithm to get the sampling probabilities at the beginning of every epoch. When sampling batches, we randomly sample with these probabilities or weights specified for each image.

In summary, unlike training the DCNN where the probability of selecting each image is 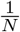, we intelligently assign probabilities to each image based on their latent space *µ* values. We then perform a weighted random selection, instead of a purely random selection, to create our training batches. Using this method, we can train the LatentNet.

### 3.5 LatentNet Training

We used three different loss functions to train the LatentNet. We continued to use binary cross-entropy of equation 1 on the sigmoid output neuron. We added two other loss functions necessary to train the VAE aspect of LatentNet. We introduced a second loss function as in equation 2 to measure the accuracy of the decoder’s reconstruction using the average pixel-wise difference. In this equation, *x* and *x*^ are the pixels of the input image and its reconstruction, respectively. In addition, we include the Kullback-Leibler divergence loss function as in equation 3 to keep the latent *µ* and *σ* close to a standard normal distribution (Kingma and Welling, 2013).

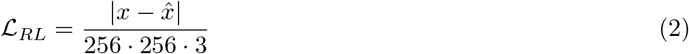

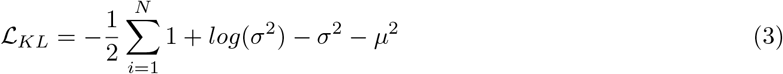

Our final loss function is summarized in Equation 4. We introduce a coefficient to ℒ_*KL*_ to scale this term down to the same level of ℒ_*B*_ and ℒ_*RL*_.

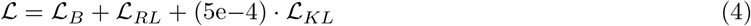

For sound comparison, the LatentNet is trained with the same hyperparameters as the DCNN with the Adam optimizer, learning rate of 5e-4, and 25 epochs.

## 4 LatentNet Evaluation

For the evaluation, we constructed both the standard DCNN and LatentNet models using TensorFlow with Keras using Python machine learning libraries. For computational efficiency, all code was executed on a NVIDIA K80 GPU.

Two datasets were utilized for this evaluation. The first set consisting of 32,542 non-Melanoma and 584 Melanoma images were acquired from the 2020 ISIC-SIIM dataset. From this dataset, a total number of 1,168 training images were used consisting of all of the 584 Melanoma images along with equal number of non-Melanoma images. The second set consisting of 1,113 Melanoma images were acquired from the ISIC 2018 archive (Tschandl et al., 2018). For LatentNet testing we used all of these melanoma images along with the remaining 31,958 non-Melanoma images from the first dataset.

### 4.1 Skin Categories

The purpose of this study is to examine whether the LatentNet is able to produce better diagnosis across all skin features and variations than the DCNN. In this section, we first develop computer vision algorithms to segregate all the 33,071 testing images into four general categories. This enables us to determine the bias in each model by comparing accuracy across these categories. The following are the categories:

1. Type I images in Fitzpatrick scale
2. Type II images in Fitzpatrick scale
3. Type III images in Fitzpatrick scale
4. Images containing visible hair

The Fitzpatrick scale is a classification system which takes into account the color of skin, tanning propensity, and general response to solar radiations (Salehinejad et al., 2018; Fitzpatrick, 1988). However, the scale is commonly used to represent the color of skin on a scale from I to VI, with type I being lightest and type VI being the darkest. In this study, we deal with the first three categories only due to lack of open-source data with images of skin from type IV to VI. the shortage of image data in these categories can be attributed to the fact that people with brown or black skin (type V and VI) are significantly less likely to get melanoma. They contain a pigment called *eumelanin* which provides extra protection from ultraviolet radiation reducing the risk of Melanoma (Davis et al., 2019). Note that the first three categories are mutually exclusive. In other words, an image can only fall into one category in Fitzpatrick scale. However, an image can simultaneously fall into the forth category and one of the first three categories.

Since the original images are not labelled with categories, we now proceed to describe two computer vision classification algorithms we developed to estimate the approximate category a dermascopic image falls into. The fist algorithm determines which one of the three Fitzpatrick scale categories it belongs. The second classification algorithm is then used to identify images with dense visible hair.

### 4.2 Fitzpatrick Scale Classifier

This algorithm is designed to place each image in the skin color category with the help of three distinct predefined image masks. First, we blur the image to remove unnecessary details accentuating the background skin color. Then the three masked images are created by keeping only the pixels that satisfy the masking logic and turning the rest to black. Finally the image is classified into the type for which the masked image has the minimum number of black pixels.

In the sample image shown in figure 7, we see that the masked image in (c) resulted in the least black pixels resulting in a Type I Fitzpatrick scale classification. This algorithm was applied to all images with and without Melanoma lesions used for LatentNet testing. Each image was stored in its respective category.

**Figure 7:**
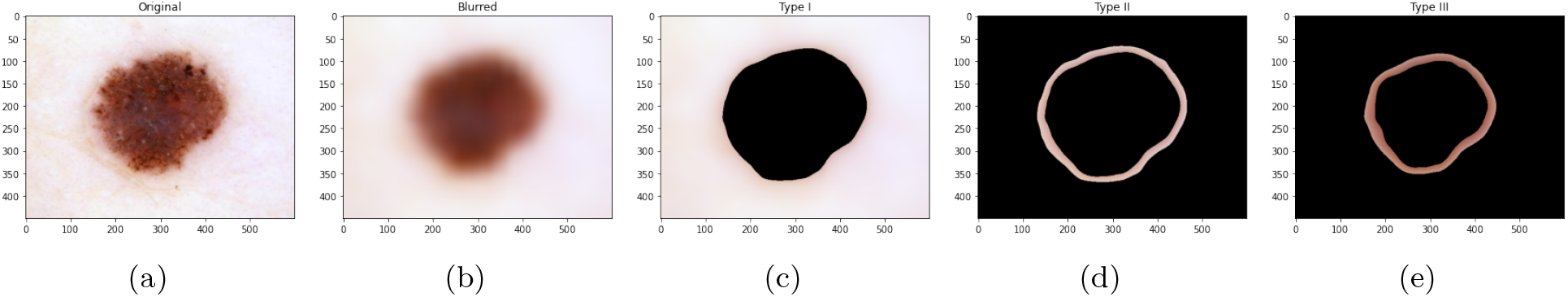
Fitzpatrick scale classification example. (a) Original Type I image. (b) The blurred image. (c-e) Masked images corresponding to Fitzpatrick Scale Types I-III masks applied to the blurred image.

### 4.3 Hair Density Classifier

A similar technique was applied to detect the images with high visible hair density. The image is first converted to gray scale so that the hair in the image is accentuated. Then, a black-hat morphological transformation is applied to filter out all but the pixels covered by hair (Adil et al., 2020). The resulting image contains binary pixel values, i.e. 0 representing background, 1 representing hair, to highlight the presence of hair. From the final binary image in Figure 8c, the hair density is calculated as the ratio of number of pixels with value one to the total number of pixels in the image. We classified an image as having high hair density if density was greater than 0.07.

**Figure 8:**
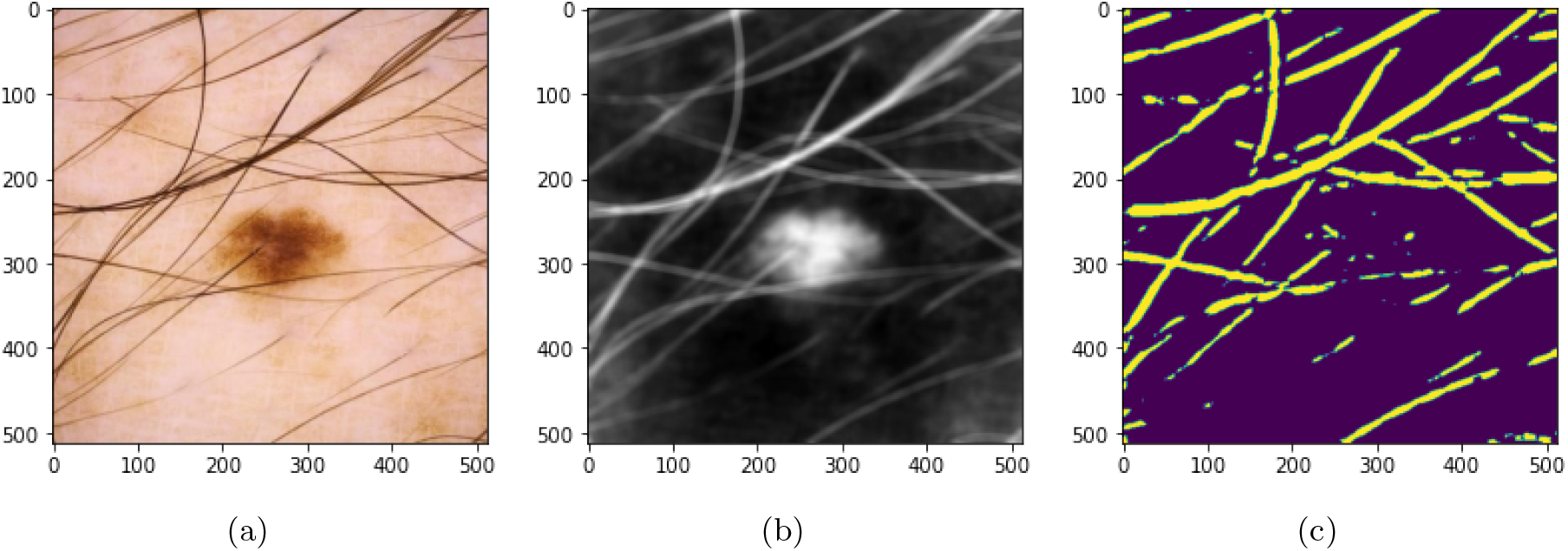
The initial image (a) is first converted to gray scale (b). Applying morphological transformation to (b) the binary image (c) is formed where the hair is identified.

After identifying hair pixels in the image, one can imagine complete removal or suppression of the hair from the image to provide more focus on the lesion itself to obtain better diagnosis accuracy. However, looking closely at the final image in figure 8c, we see that certain pixels have been highlighted that are not part of actual hair. If we suppress these pixels, we may instead be removing crucial details contributing to the diagnosis in the particular image. The purpose of this algorithm is to calculate the approximate hair density for classification of the test image. Determination of exact hair amount is not relevant for the purpose.

Sample test images from the four generated categories are shown in figure 9.

**Figure 9:**
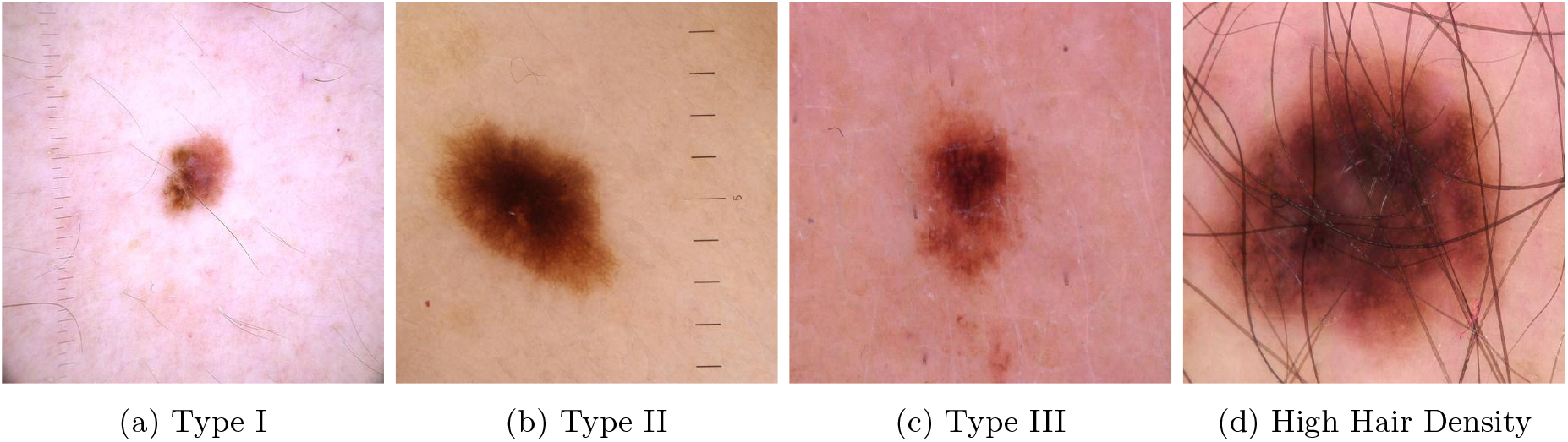
Sample images from the four test categories.

## 5 LatentNet Performance

Once the four categories of test sample datasets were created, the DCNN and LatentNet were trained and tested on the dataset of each category to collect the accuracy and the AUC scores. The above process of training and testing was repeated ten times to determine average accuracy and AUC scores for the categories.

As shown in figure 10, LatentNet showed significantly higher diagnostic accuracy in all categories except the Type I category. In the Type I group, the DCNN performed marginally higher than the LatentNet (90.41% and 89.52%, respectively) suggesting that there is Type I accuracy remained the nearly the same under LatentNet.

**Figure 10:**
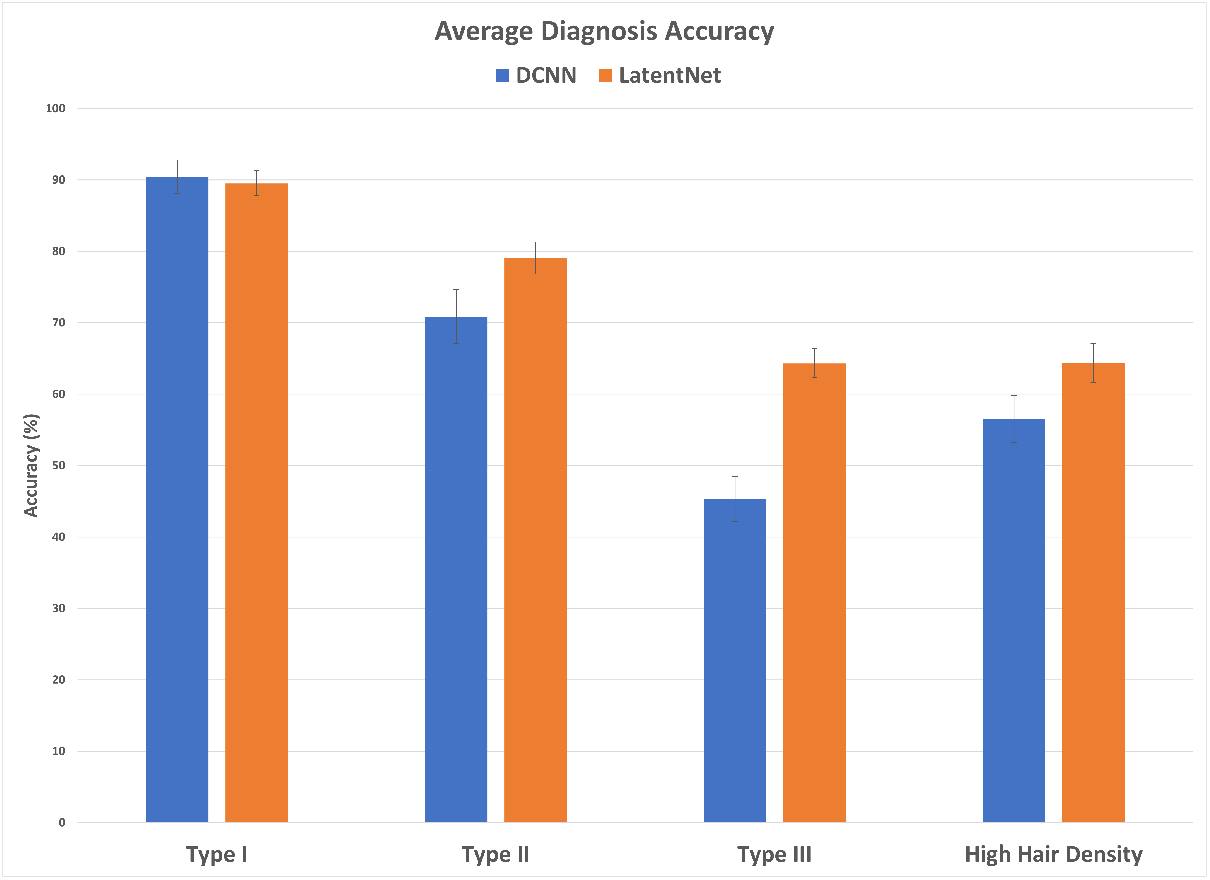
Comparison of average accuracy for each four validation categories.

We also conducted a 2-way analysis of variance (ANOVA) test to assess if the observed accuracy improvements are statistically significant. As shown in figure 11, we observe statistical significance (*p* < 0.05) in all three groups from the ANOVA. The significant difference in the model group strongly suggests that one model (LatentNet) shows better overall performance over the other model (DCNN). The *p*-value of the category group suggests that the average performance across categories differ significantly. Finally, the interaction between model and category suggests that the choice of the model makes a significant difference in the accuracy for a given category. Since the *p* value of the interaction term was less than 0.05, we conducted Tukey’s honestly significance difference (HSD) post-hoc test to determine one or more specific categories where the choice of model made a significant difference in diagnosis accuracy (Abdi and Williams, 2010).

**Figure 11:**
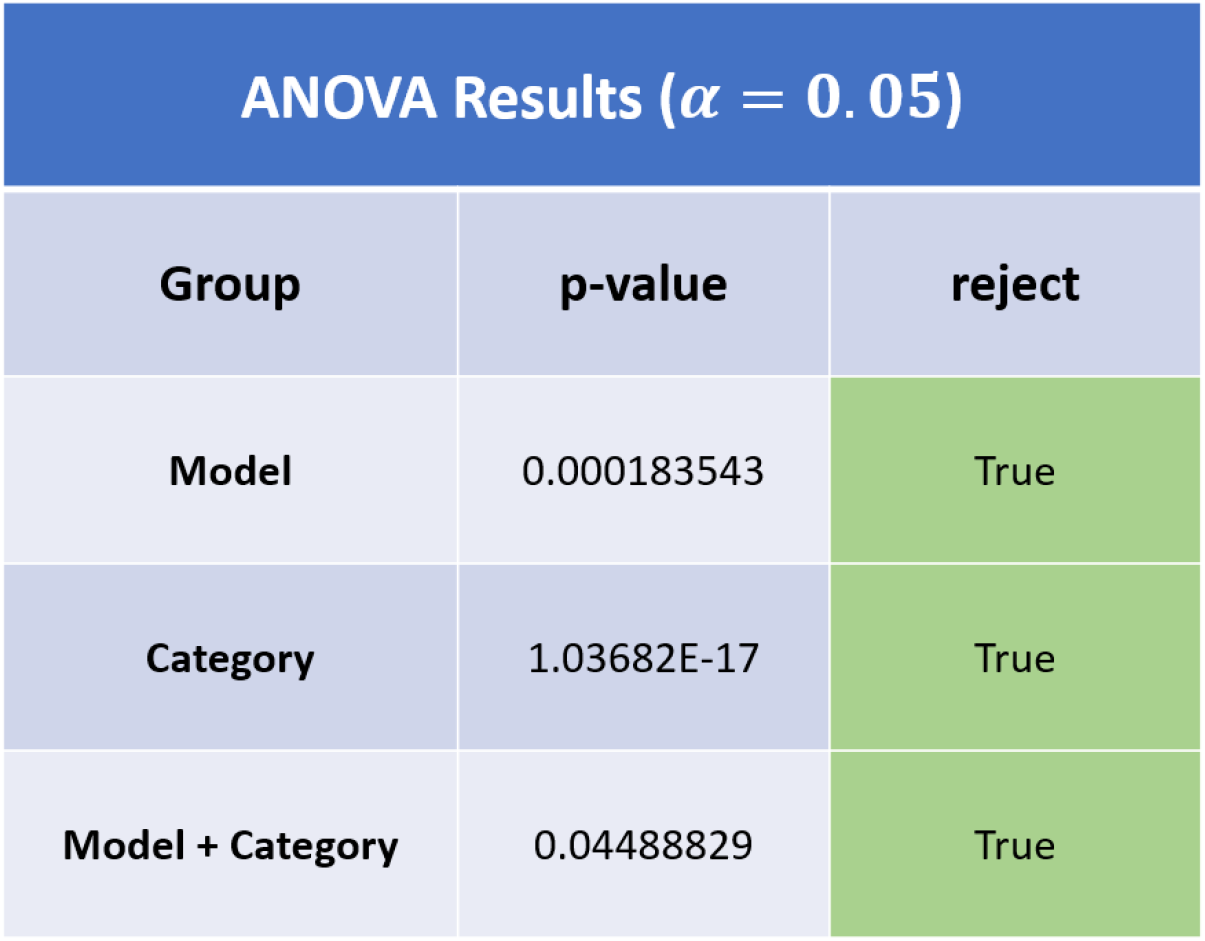
2-way ANOVA results.

Tukey’s HSD results are shown in Figure 12. We observe that the LatentNet performed significantly better than a DCNN on images that lie approximately in the Type III Fitzpatrick scale category. Although there is noticeable improvement in Type II and images with hair density as depicted in Figure 10, they are not statistically significant (*p* > 0.05). Although in the Type I category, DCNN showed trifle better accuracy, Tukey’s HSD result suggests that the accuracy difference is not significant. From the ANOVA and Tukey’s HSD tests, we can conclude that the LatentNet displayed optimal overall performance, and there is strong evidence that it indeed improves the diagnosis accuracy significantly in the Type III category.

**Figure 12:**
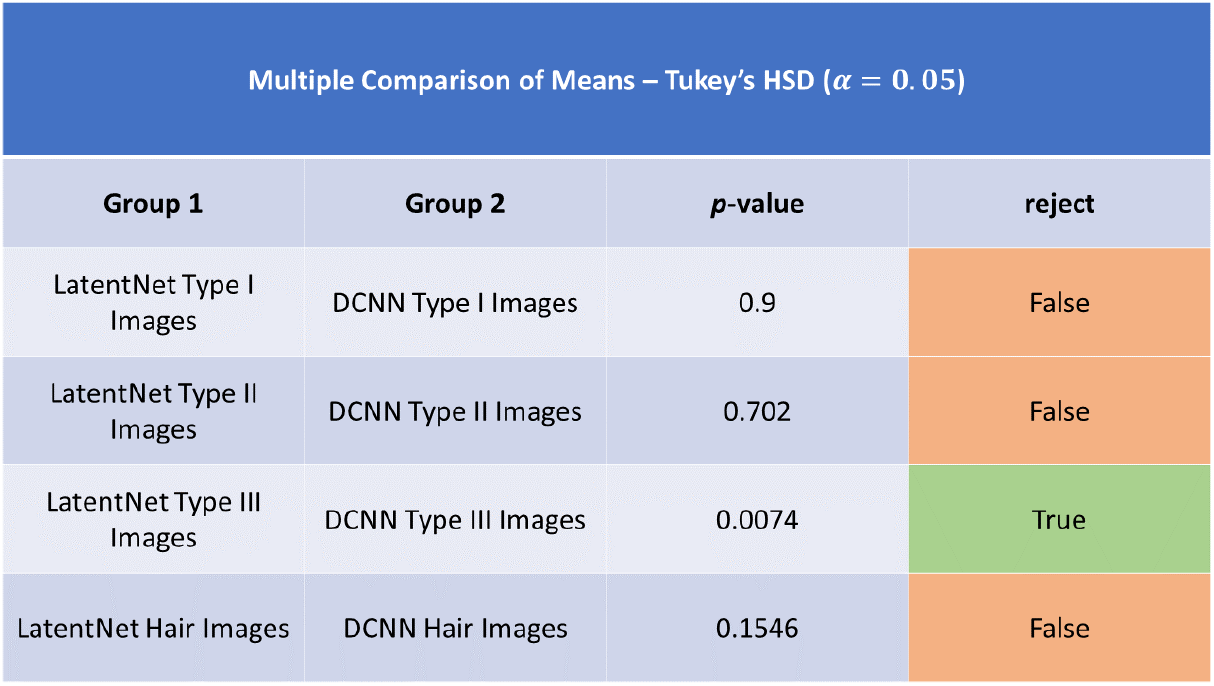
Results of the Tukey’s HSD post-hoc test.

The LatentNet out-performed the DCNN in all categories except the Type I. We investigated this category further by looking at the confusion matrix as shown in Figure 13 of one particular run. Although LatentNet model performed marginally worse in the overall accuracy, the number of false positives, i.e. Melanoma cases are predicted as No-Melanoma, is significantly lower in the LatentNet (18 in LatentNet, 47 in DCNN). False positives are perhaps the most dangerous diagnostic error because the cancer goes undetected and may eventually spread. Though the false positives reduced in the LatentNet, the slight decrease in overall performance may be attributed to disproportionately large number of No-Melanoma test images.

**Figure 13:**
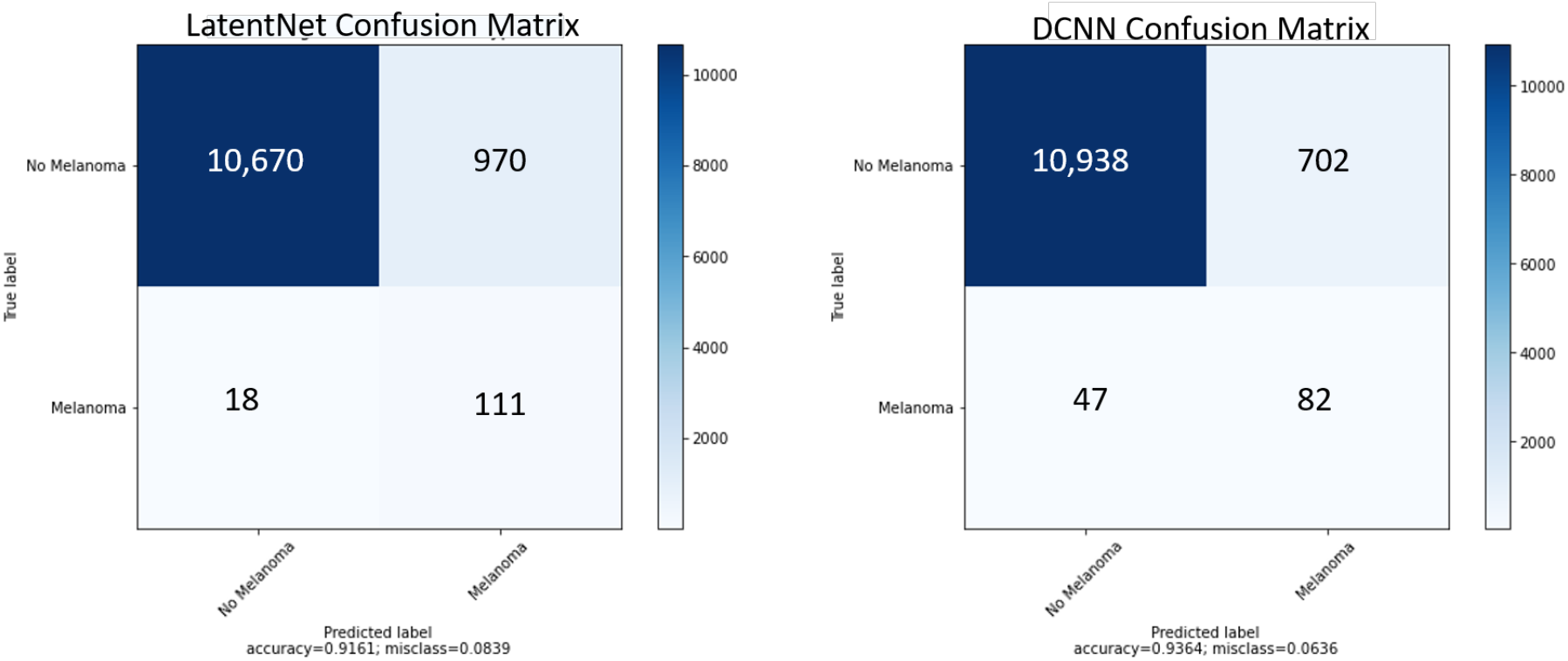
Confusion matrix for LatentNet and DCNN with Type I testing images.

**Figure 14:**
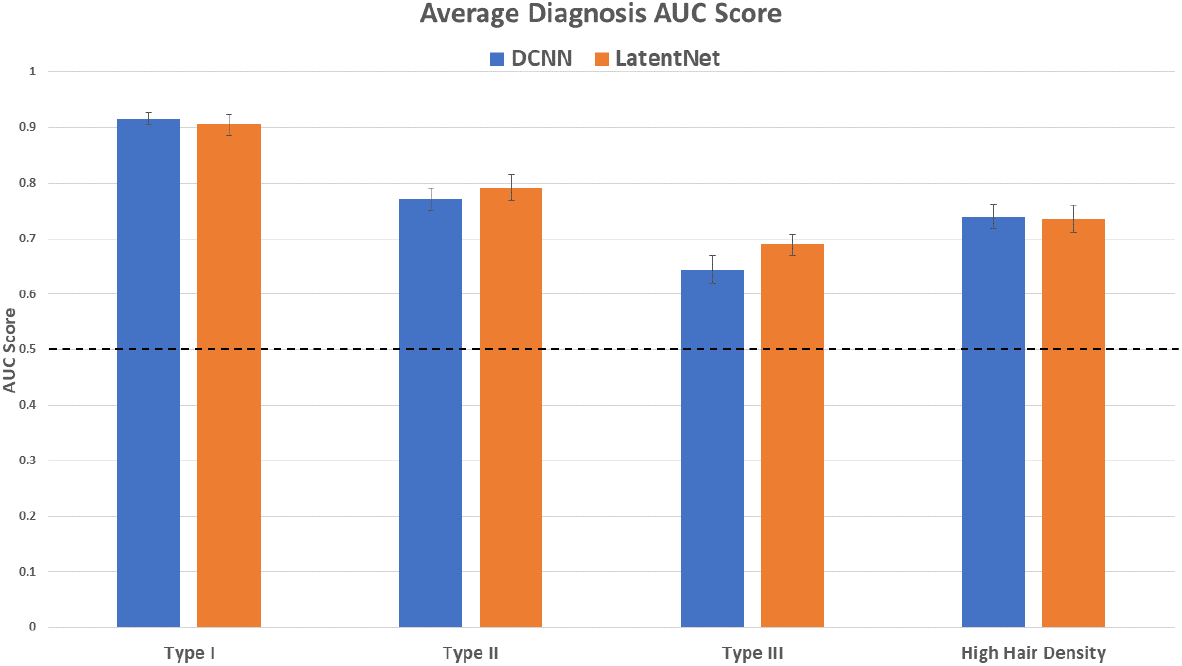
Average AUC scores over the 10 runs. Dotted line at 0.5.

The large imbalance in the Melanoma images in each category is perhaps best explained by the fact that the disease is extremely rare but also fatal compared to ordinary skin lesions. Under these circumstances, measuring the AUC score helps us ensure that the LatentNet is not merely predicting No-Melanoma always. This can potentially produce a false sense of high accuracy as well.

The AUC score tells us the ability of our model to discriminate between the two diagnostic classes, viz. Melanoma and No-Melanoma. By definition the AUC score is 0.5 if a model always predicts the same class. It is important to notice that both the models predict with AUC score higher than 0.5 for all categories. The LatentNet shows better discrimination among classes for Type II and III images. However, the DCNN shows marginally higher AUC scores for Type I and high hair density images.

Especially for Type I category, LatentNet fares slightly worse than the DCNN model taking both accuracy and AUC score into consideration. This may be caused by over representation of Type I images in the original dataset. In such a scenario, each batch of training images for the DCNN, which relies on pure random sampling, will be skewed towards Type I images. As a result, the DCNN will overfit and will have a propensity towards learning features present only in Type I images. However, the LatentNet detects light background skin color in Type 1 images and intelligently reduces the sampling probability for that particular image. Therefore, the likelihood of over-fitting to such images is significantly lower in the LatentNet.

## 6 Conclusions

This study presents a fully automated diagnosis of Melanoma while addressing inherent dataset bias. We develop a model called LatentNet which is different from existing state-of-the-art models by virtue of its ability to intelligently reduce bias in each training batch. The algorithm does not need an explicitly defined category to reduce bias in, instead, it samples based on its learned latent structure. The latent structure is learned in a completely unsupervised process through a variational autoencoder (VAE), not needing specific labels denoting the under-represented images. However, we train the subsequent classifier phase of the model using a supervised algorithm effectively following an overall semi-supervised training process.

We extensively tested the LatentNet model to evaluate performance gain relative to the baseline DCNN model on four separate test image categories. We discovered that our proposed approach increased the overall diagnostic accuracy in dermascopic images with darker background skin color, corresponding to images in Type II and III Fitzpatrick scales. We also observed higher accuracy using the LatentNet in images with high visible hair density.

The LatentNet model, however, experienced marginal decrease in overall accuracy among patients with light skin color in the Type I Fitzpatrick Scale category. Upon close examination of a representative confusion matrix, it seems that the LatenNet significantly improves the accuracy of the images having Melanoma. There is a slight degradation of accuracy for the images having no Melanoma. Overall accuracy suffers since there are a lot more non-Melanoma images compared to the Melanoma images in the test set.

In the future, we plan to evaluate the new model with larger set of Melanoma images to counter lopsided representation of non Melanoma images. We will also explore the use of an ensemble model which would produce an output taking advantage of both the standard DCNN and the LatentNet model intelligence. This may improve the performance for all image categories. An alternative approach to first determine the category of the input image (e.g. Type I, Type II or Type III) first and then use the model that works best for the given category.

For this research, we built a simple 5 layer DCNN model. This was partially done because we joined the variational autoencoder with the classification model. In other words, the classification portion of our model was acting as the encoder of a VAE. Since this was the encoder, we had to write a transposed version of the model as the decoder. This limited us to keeping a basic sequential model. However, it would be beneficial to utilize transfer learning and implement larger model such as the ResNet50 or VGG16 (He et al., 2016; Simonyan and Zisserman, 2014). To implement this, we would have to separate the classification model from the VAE. More specifically, at every epoch, we would first train a plain VAE and compute the sampling probablities for each training image. Then, we would perform our intelligent selection to form a batch which we pass into the larger classification model (e.g. ResNet50, VGG16, etc.). This way we would be able to detect more complex features from the input images.

In summary, we built a new machine learning model that addresses the issue of racial bias in a dataset. We demonstrated that this unbiased model performed significantly better than the commonly used DCNN. This elevates the prospect of improved early Melanoma diagnosis and better survival chance for a vastly diverse world population.

## Data Availability

The data that supports the findings of this study are openly available from the International Skin Imaging Collaboration archives.

https://challenge2020.isic-archive.com/

https://challenge2018.isic-archive.com/

